# Predicting clozapine initiation among patients with schizophrenia via machine learning trained on electronic health record data

**DOI:** 10.64898/2026.04.17.26351083

**Authors:** Erik Perfalk, Jakob Grøhn Damgaard, Andreas Aalkjær Danielsen, Søren Dinesen Østergaard

## Abstract

**Background and Hypothesis:** Clozapine is the only medication with proven efficacy for treatment-resistant schizophrenia, yet many patients experience delays of several years before initiation. Our aim was to develop and validate a dynamic prediction model for clozapine initiation among patients with schizophrenia trained solely on electronic health record (EHR) data from routine clinical practice.

**Study Design:** EHR data from all adults (≥ 18 years) with a schizophrenia (ICD10: F20) or schizoaffective disorder (ICD10: F25) diagnosis who had been in contact with the Psychiatric Services of the Central Denmark Region between 1 January 2013 and 1 June 2024 were retrieved. 179 structured predictors were engineered (covering, e.g.,diagnoses, medications, coercive measures) and 750 predictors derived from clinical notes. At every psychiatric hospital visit, we predicted if an incident clozapine prescription occured within the next 365 days. XGBoost and logistic regression models were trained on 85% of the data with 5-fold stratified cross-validation. Performance was evaluated on the remaining 15% of the data (held out) using the area under the receiver operating characteristic curve (AUROC).

**Study Results:** The training/test set comprised of 194,234/35,527 hospital visits, distributed on 4928/878 unique patients. In the test set, the best XGBoost model achieved an AUROC of 0.81, sensitivity of 32%, positive predictive value of 23% at a 7.5% predicted positive rate.

**Conclusions:** A dynamic prediction model based solely on EHR data predicts clozapine initiation with high discrimination. If implemented as a clinical decision support tool, this model may guide clinicians towards more timely initiation of clozapine treatment.

## Introduction

Clozapine is the primary treatment for patients with treatment-resistant schizophrenia.^1^ However, studies report delays of up to 48 months before clozapine initiation, despite clear indication and the known association between this delay and worsened prognosis.^2^ Prediction of treatment resistance could allow for timely clozapine use, improving outcomes and reducing long-term burden.^3^

Previous studies have aimed at predicting treatment resistance or clozapine initiation using clinical data.^4–6^ However, these studies employed a static prediction approach, generating risk estimates only at the time of diagnosis or initiation of a treatment course (i.e., enrollment in early intervention services). While predictions timed this way can provide clinicians with very early alerts about potential treatment resistance, they fail to incorporate updated clinical information that becomes available over time, thereby overlooking dynamic changes in a patient’s risk profile.^7^ In contrast, a dynamic, “landmark” modelling approach (e.g., predicting at every outpatient contact) can incorporate evolving clinical data, potentially improve predictive performance, and enable more timely and targeted risk prediction.

To our knowledge, no previous studies have employed a landmark approach to predict clozapine initiation. We have previously shown that such dynamic prediction can be effectively applied to other clinically outcomes for patients with mental illness.^8–13^ Dynamic prediction modelling also aligns more naturally with clinical workflows. Therefore, the aim of this study was to develop and validate a dynamic machine learning–based prediction model for clozapine initiation using only routinely collected electronic health record (EHR) data, the latter facilitating potential clinical implementation.

## Methods

An illustration of the methods used in this study is shown in Figure 1.

**Figure 1:**
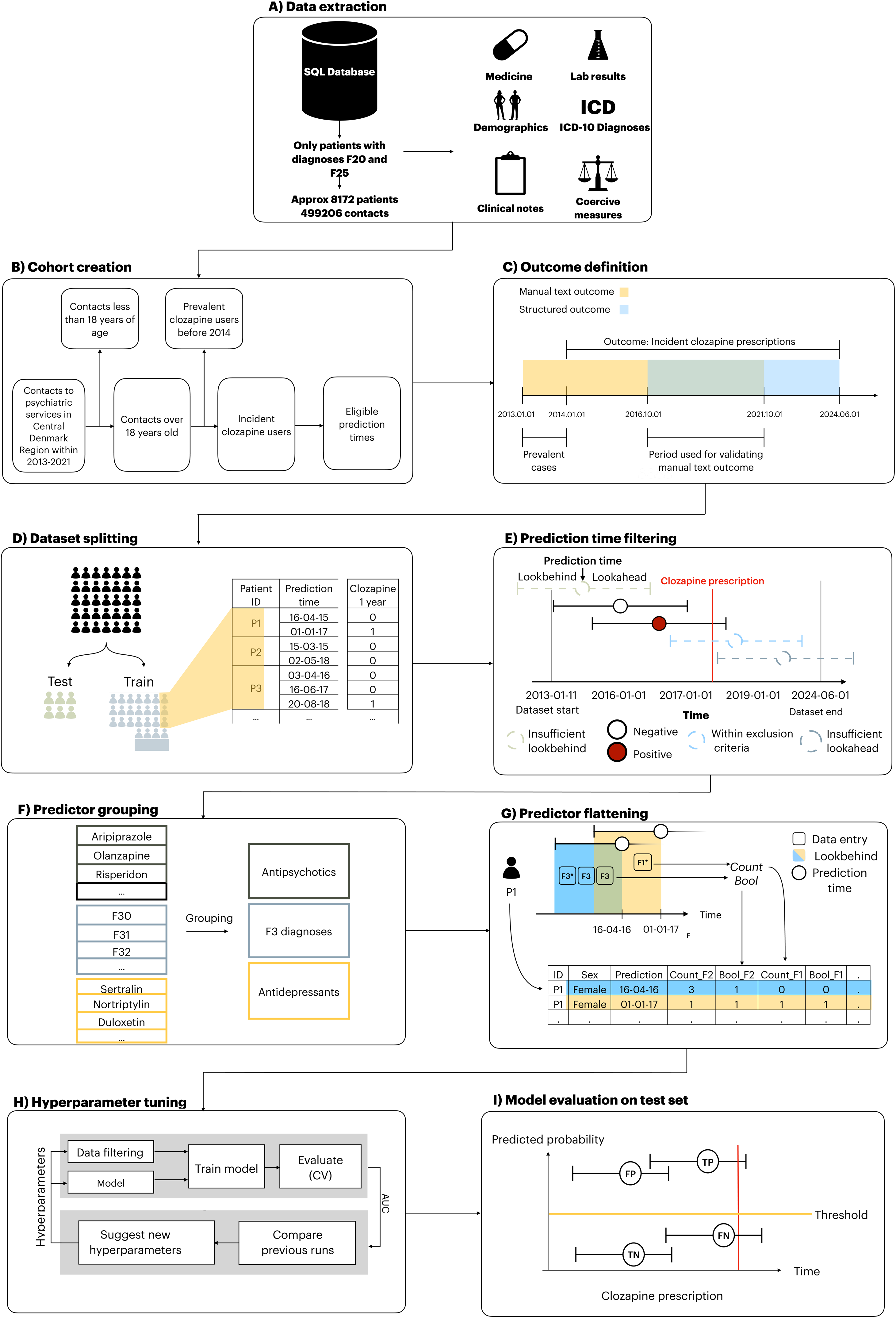
Model development (Extraction of data, cohort creation, outcome definition, dataset splitting, prediction time filtering, specification of predictors and flattening, model training, testing and evaluation) Figure was modified to this project based on Bernstorff et al (Bernstorff et al., 2024). F1 and F3: ICD-10 diagnoses within the group of diagnoses included in F1 chapter and F3 chapter. CV: Cross-validation. TP: True positive. FP: False positive. TN: True negative, FN: False negative

### Reporting guidelines

This study adhered to the reporting guidelines of TRIPOD+AI.^14^ The TRIPOD+AI checklist is available in the Supplementary Material (Supplementary Table 8).

### Data source

The study is based on routine electronic health record (EHR) data from the Psychiatric Services of the Central Denmark Region. The Central Denmark Region is one of five Danish Regions and has a catchment area of approximately 1.4 million people. The dataset contains timestamped records of all contacts with public hospitals in the Central Denmark Region, including both psychiatric and general medical hospital services. Given Denmark’s universal healthcare system, the vast majority of hospital contacts occur within public hospitals (there are no private psychiatric hospitals in Denmark) and, are therefore captured in this dataset.^15–17^

### Cohort definition

The cohort consisted of all adult patients (aged 18 or above) with a schizophrenia (ICD-10: F20) or a schizoaffective disorder (ICD-10: F25) diagnosis and at least one contact to the Psychiatric Services of the Central Denmark Region in the time period from January 1, 2013- June 1, 2024 (see Figure 1A). We included schizoaffective disorder as its ICD-10 operationalization essentially requires presence of the full schizophrenia syndrome and because clozapine is also used in treatment-resistant schizoaffective disorder.^18,19^ (see Figure 1B).

### Outcome definition

The outcome was defined as the first clozapine prescription. Due to instability in the prescription data prior to October 2016, we only used prescription data for outcome definition from October 2016 and onwards (see figure 1C). To enable inclusion of data from 2013- September 2016, we performed manual classification of clozapine initiation using free-text clinical notes for that period. First, we searched for mentions of ”clozapine” and ”leponex” (the trade name for clozapine in Denmark) in the clinical notes in this period. To account for potential misspellings, we calculated a similarity ratio^20^ using the Python package rapidfuzz v. 3.6.1. Subsequently, these notes were read by EP and if clozapine was initiated, this was labelled and timestamped. Cases of uncertainty were discussed with AAD and SDO. One of the major groups of uncertain cases was patients who had previously received clozapine (for example between 2005–2008) but were not receiving it during the period covered by the study. Since the goal was to obtain data comparable to the prescription data, we did not exclude these as prevalent cases, as they would appear as incident cases in the period where prescription data were used to define clozapine initiation. To ensure the validity of the manual text validation process, we used the method described above to also manually classify clozapine initiation based on clinical notes from October 1, 2016 to November 22, 2021 and compared it to the clozapine prescription data. See Supplementary Table 1 for further details on handling of uncertain cases.

### Dataset splitting

The patients were randomly split into a training (85%) and a test (15%) set (see Figure 1D). The test set was held out until the final stage of model evaluation, where no additional changes were made to the model developed in the training phase.

### Prediction times and exclusion criteria

Prediction times were defined as all outpatient- and start of inpatient psychiatric hospital contacts. To remove prevalent clozapine users, patients registered with a prescription for clozapine or a plasma clozapine laboratory test between 1 January 2013 and 31 December 2013 were excluded. Prediction times were excluded if they lacked a sufficient lookbehind window (for extracting predictor variables) or a sufficient lookahead window (for determining outcomes). The definitions of the lookbehind and lookahead windows are provided below (see also Figure 1E).

### Lookahead window

The lookahead window (the period following the prediction time in which the outcome could occur) was set to 365 or 730 days. Hence, all prediction times for which a clozapine prescription occurred within 365 or 730 days were deemed to be positive outcomes (see Figure 1E).

### Predictor engineering and lookbehind window

A complete list of the predictors (n= 929) and their definitions is available in Supplementary Table 2. Predictors were selected based on literature on characteristics associated with clozapine treatment^21^ and supplemented with clinical domain knowledge. The predictors fall into ten categories: age and sex, hospital contacts, psychiatric diagnoses, administered medications, laboratory results, coercive measures, psychometric rating scales, suicide risk assessment, electroconvulsive therapy (ECT) and free text predictors from EHR clinical notes (extracted using term frequency-inverse document frequency^22^ – see below for details). Hospital contacts cover both admissions and outpatient contacts along with registered diagnoses for these contacts. Diagnostic predictors were the psychiatric subchapters (F0-F9 with F2 excluded) from the International Classification of Disease (ICD-10)^23^ with bipolar disorder (F30-F31) and cluster B-personality disorders (F60.2-F60.4 (dissocial-, borderline- and histrionic personality disorder) as additional separate predictors. Medication predictors were based on structured Anatomical Therapeutic Chemical (ATC) classification system codes^24^ and grouped as follows (Figure 1F): antipsychotics, first generation antipsychotics, second generation antipsychotics, depot antipsychotics, antidepressants, anxiolytics, hypnotics/sedatives, stimulants, analgesics, and drugs for treatment of alcohol abstinence/opioid dependence. Lithium and olanzapine were additionally included as individual predictors. Laboratory-based predictors included plasma analysis of antipsychotics, antidepressants, paracetamol, and ethanol. Coercive measures encompassed involuntary admissions and treatments, manual restraint, chemical restraint, and mechanical restraint (belt). Psychometric rating scales included scores from the Brøset violence checklist (a 6-item clinician-rated checklist assessing acute risk of violent behaviour)^25^ and suicide risk assessment (a scoring system used in the Central Denmark Region with the following risk levels: 1 - no increased risk, 2 - increased risk, and 3 - acutely increased risk)). ECT was included as a binary predictor (yes/no). Free text predictors were derived from 17 clinical note categories from the EHR considered both clinically informative and temporally stable, e.g., “Subjective Mental State” and “Current Objective Mental State” (see Supplementary Table 3 for the full list).^17^ We applied TF-IDF^22^ to create weighted representations of the clinical notes . First, the text was preprocessed by lower-casing all words and removing stop words and symbols. Subsequently, the model generated all uni- and bi-grams. Secondly, uni- and bi-grams with a document frequency above 90% by were removed (due to assumed low predictive value). Lastly, the top 750 uni- or bigrams with highest document frequency within the 90% threshold were included in the model. This model was used to vectorize all individual notes in the dataset. For each prediction time, notes from the last 180 days were extracted and their TF-IDF vectors were averaged together to create a single TF-IDF embedding with 750 values (based on the 750 top uni- or bigrams) which were used as predictors.

Predictors were constructed by aggregating the values for the variable of interest within a specified lookbehind window (30, 180 and 365 days leading up to a prediction time) using different predictor aggregation functions (mean, max, bool, etc.). The specific aggregations methods for each variable can be found in Supplementary Table 2. Data processing was performed using the timeseriesflattener v2.0.1 package (Figure 1G) ^26^. If a predictor was absent in the lookbehind period, it was coded as “missing”. However, these instances do not indicate missing values in the traditional sense, as they stem from a true absence of clinical records, rather than, e.g., a missed visit in a clinical trial. As this mirrors routine clinical practice, such cases were retained to ensure consistency with real-world availability in a potential implementation setting.

### Predictor sets

A priori, we pre-defined four predictor sets to investigate the predictive performance balanced against the number of predictors:

- Demographics + count of antipsychotics used (n=5)
- Demographics + structured predictors, i.e., all predictors not based on EHR text (n=179)
- Demographics + TF-IDF, i.e., predictors from EHR text (752 predictors)
- Demographics + structured predictors + TF-IDF (929 predictors)

### Hyperparameter tuning and model training

Two machine learning approaches were trained: XGBoost and elastic net-regularized logistic regression (using scikit learn version 1.2.1).^22^ XGBoost was selected due to the strong predictive performance of gradient boosting techniques on structured data, their computational efficiency, and inherent ability to accommodate missing values.^27,28^ Elastic net regularized logistic regression served as a benchmark model.^29,30^ 5-fold stratified cross-validation was employed for training with no patient occurring in more than one fold. Hyperparameter optimization (see Supplementary Table 4 for details) was conducted across 150 trials with the aim of maximizing the area under the receiver operating characteristic curve (AUROC). Optimization was performed using the tree-structured parzen estimator method in Optuna v2.10.1.33 (see Figure 1H).^31^ All analyses were carried out in Python (version 3.10.9).

### Model evaluation on test data

All model configurations were evaluated on the test set (see Figure 1I). In addition to AUROC, we calculated the sensitivity, specificity, positive predictive value (PPV), negative predictive value (NPV), and F1 at predicted positive rates (PPR) of 1%, 2%, 3%, 4%, 5%, 7,5% 10%, 20% and 50%, respectively. The predicted positive rate (threshold) represents the proportion of all prediction times classified as positive. To assess robustness, the best performing model was further evaluated across subgroups defined by sex-, age-, calendar month, and days until outcome.

Additionally, model calibration was assessed with calibration curves accompanied by distribution plots of the predicted probabilities for the best XGBoost and logistic regression models. Clinical utility was evaluated by decision curve analysis.^32^ Plots were truncated at an upper threshold of 0.20, corresponding to a one-in-five probability of receiving a clozapine prescription within one year should nothing change, and it is unlikely that risk thresholds greater than this would be tolerated. Net benefit was defined as the additional proportion of true cases identified through the use of the models without a corresponding increase in false positives.

### Estimation of predictor importance

To examine which predictors contributed most to model predictions, we computed measures of predictor importance. For the XGBoost models, importance was estimated using information gain.^33^ In this context, the gain associated with a predictor is calculated as the average improvement in loss when generalizing to the training data accomplished by the predictor across all node splits in the model that handle the predictor. For logistic regression models, predictor importance was assessed using standardized regression coefficients.^34^ These coefficients represent the change in log-odds for a one standard deviation increase in the predictor values. The magnitude of each coefficient therefore indicates the strength of the association between the predictor and the outcome while adjusting for all the other predictors, enabling comparison of relative importance across predictors. Notably, coefficients from logistic regression are directional, indicating whether higher predictor values increase or decrease the probability of a positive outcome. In contrast, information gain from XGBoost reflects overall contribution to predictive performance and does not convey the direction of the association.

### Post hoc analysis

We performed cross-validated model training after removing “clozapine” and “leponex” as text predictors to evaluate the impact of their exclusion. This was done to assess the risk that the model primarily captured clinicians’ prior considerations regarding clozapine and made predictions based on that information.

### Ethics

The study was approved by the Legal Office of the Central Denmark Region in accordance with the Danish Health Care Act §46, Section 2 (reg, no. 1-45-70-61-24). The Danish Committee Act exempts studies based only on EHR data from ethical review board assessment (waiver for this project: 1-10-72-103-24). Handling and storage of data complied with the European Union General Data Protection Regulation. The project is registered on the list of research projects having the Central Denmark Region as data steward (reg, no. 1-16-02-398-24). There was no patient -nor public involvement in this study.

## Results

### Cohort

A flowchart illustrating the definition of the cohort is shown in Supplementary Figure 1. The dataset eligible for prediction consisted of 269,529 psychiatric outpatient hospital contacts (prediction times) distributed on 6303 unique patients. After randomization, the training set consisted of 225,643 prediction times distributed on 5235 unique patients and test set 46,701 prediction times distributed on 1068 unique patients.

Table 1 lists clinical and demographic patient data for the prediction times included in the training and evaluation of the main model. The main model included predictors with a lookbehind window of up to 365 days. After removing all prediction times where the lookbehind or lookahead windows extended beyond the available data for a patient, a total of 229,761 (training set n==194,234, test set n=35,527) prediction times remained. These prediction times were distributed across 5806 (training set n = 4928, test set n = 878) unique patients (with the following demographics: 43.5% females (training set = 43.6% and test set = 43.3%), median age [Q1,Q3] = 32.1 years [24.9,44.1] (training set = 32.2 [24.9,44.1] and test set = 31.9 [24.8,44.1])). A total of 9400 (training set n=7959, test set n=1441) of the outpatient contacts were followed by clozapine initiation within 365 days (positive outcome), distributed across 589 (training set= 506, test set=83) unique patients (clozapine initiation could be included in multiple positive outcomes as a patient could have multiple outpatient contacts (prediction times) in the 365 days prior to clozapine initiation.

**Table 1.**
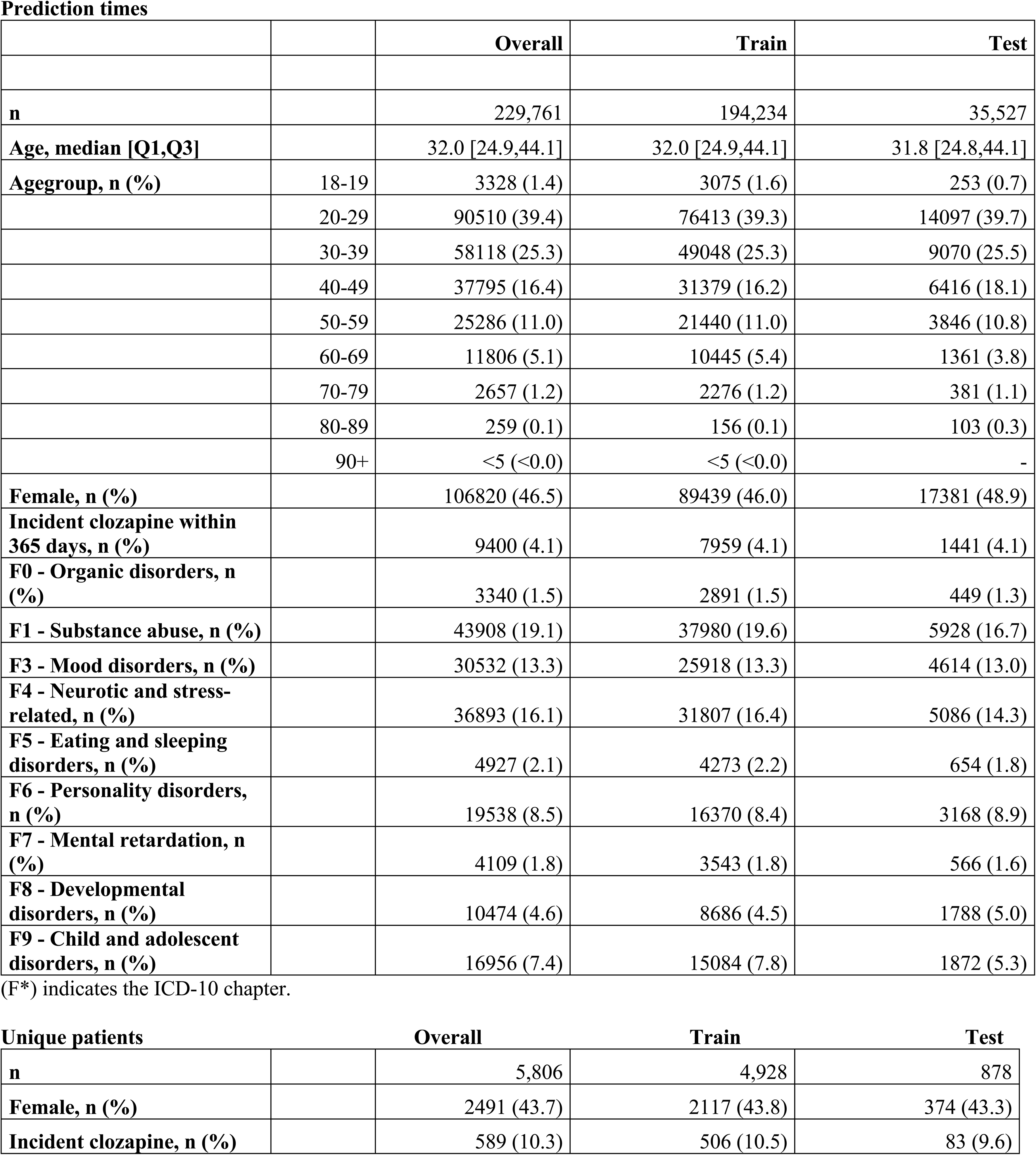
Descriptive statistics for best model (365 days lookahead).

### Manual classification of clozapine initiation based on text from clinical notes

The text similarity (fuzz ratio > 70) search rendered 94,092 clinical notes distributed on 3846 unique patients with a median of 7 notes/patient. The manual text classification added 888 outcomes of which 634 where in the washout period from 2013-2014 (prevalent users). Comparison of manually classified clozapine initiation with that defined by prescription data for the period from October 2016 to November 2021 showed high agreement. Specifically, the median difference in the date of clozapine initiation was 0 days and 85% of the dates of initiation were within 10± days (see Supplementary Figure 2 and 3).

### Model evaluation

Following cross-validation on the training set, XGBoost (Best model: Full predictor set – 365 days lookahead AUROC = 0.80) outperformed logistic regression (Best model: Full predictor set – 365 days lookahead AUROC = 0.77) across all model variations. The models with 365 days lookahead outperformed models with 730 days lookahead. Models trained using only the TF-IDF predictors and demographics showed similar performance to those using the full set of predictors, while models with only structured predictors had relatively lower AUROC (see Table 2). The hyperparameters used for the best XGBoost and elastic net models are listed in Supplementary Table 5. When evaluated on the test set, all models maintained the same level of performance as in the training phase (Best model AUROC: XGBoost = 0.81, Logistic regression = 0.78) (see Table 2). Figure 2 shows the performance of the best models, with robustness measures in Supplementary Figures 4 and 5.

**Figure 2:**
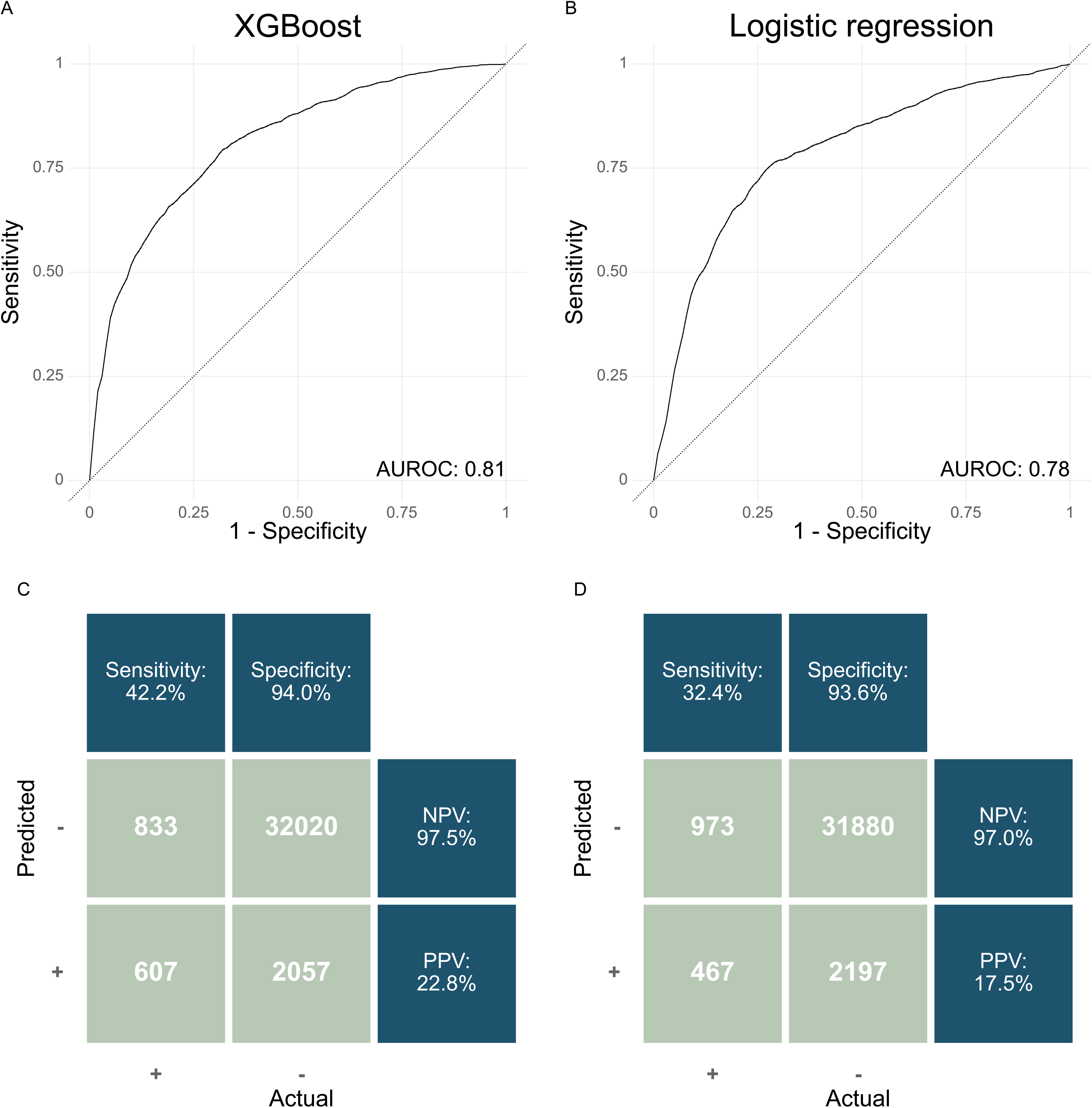
Model performance of the XGBoost and logistic regression on the test set. A & B) Receiver operating characteristics curve. AUROC= Area under the receiver operating characteristics curve. C & D) Confusion matrix. PPR: Positive predictive rate. NPV: Negative predictive value. The decision threshold is defined based on a predicted positive rate of 7.5%. More results from the models are presented in Table 3.

**Table 2.** Model performance on training and test set for XGBoost and Logistic regression models trained on different subsets with different lookaheads (2A & 2B).

| **2A. 365 days lookahead** |  | **Training set** |  | **Test set** |
| --- | --- | --- | --- | --- |
| **PREDICTOR SET** | **Number of predictors** | **Mean Internal AUROC** | **5-fold internal AUROC range** | **AUROC** |
| **XGboost** |  |  |  |  |
| Full predictor set | 929 | 0.80 | [0.78:0.82] | 0.81 |
| Only structured | 179 | 0.72 | [0.64:0.78] | 0.73 |
| Only TF-IDF | 752 | 0.79 | [0.77:0.81] | 0.79 |
| Unique antipsychotics | 5 | 0.70 | [0.62:0.77] | 0.725 |
| **Logistic Regression** |  |  |  |  |
| Full predictor set | 929 | 0.77 | [0.74:0.80] | 0.78 |
| Only structured | 179 | 0.71 | [0.67:0.76] | 0.73 |
| Only TF-IDF | 752 | 0.74 | [0.72:0.76] | 0.76 |
| Unique antipsychotics | 5 | 0.71 | [0.65:0.76] | 0.731 |
| **2B. 730 days lookahead** |  |  |  |  |
| **PREDICTOR SET** | **Number of predictors** | **Training set** |  | **Test set** |
|  |  | **Internal AUROC** | **5-fold internal AUROC range** | **AUROC** |
| **XGBoost** |  |  |  |  |
| Full predictor set | 929 | 0.78 | [0.76:0.80] | 0.78 |
| Only structured | 179 | 0.71 | [0.68:0.74] | 0.75 |
| Only TF-IDF | 752 | 0.77 | [0.75:0.80] | 0.76 |
| Unique antipsychotics | 5 | 0.70 | [0.66:0.74] | 0.71 |
| **Logistic Regression** |  |  |  |  |
| Full predictor set | 929 | 0.75 | [0.74:0.78] | 0.77 |
| Only structured | 179 | 0.71 | [0.68:0.74] | 0.72 |
| Only TF-IDF | 752 | 0.75 | [0.72:0.76] | 0.74 |
| Unique antipsychotics | 5 | 0.70 | [0.68:0.74] | 0.70 |

All models included demographics (Age/Sex). Details on predictor descriptions can be found in Supplemental Table 1.

**Table 3.**
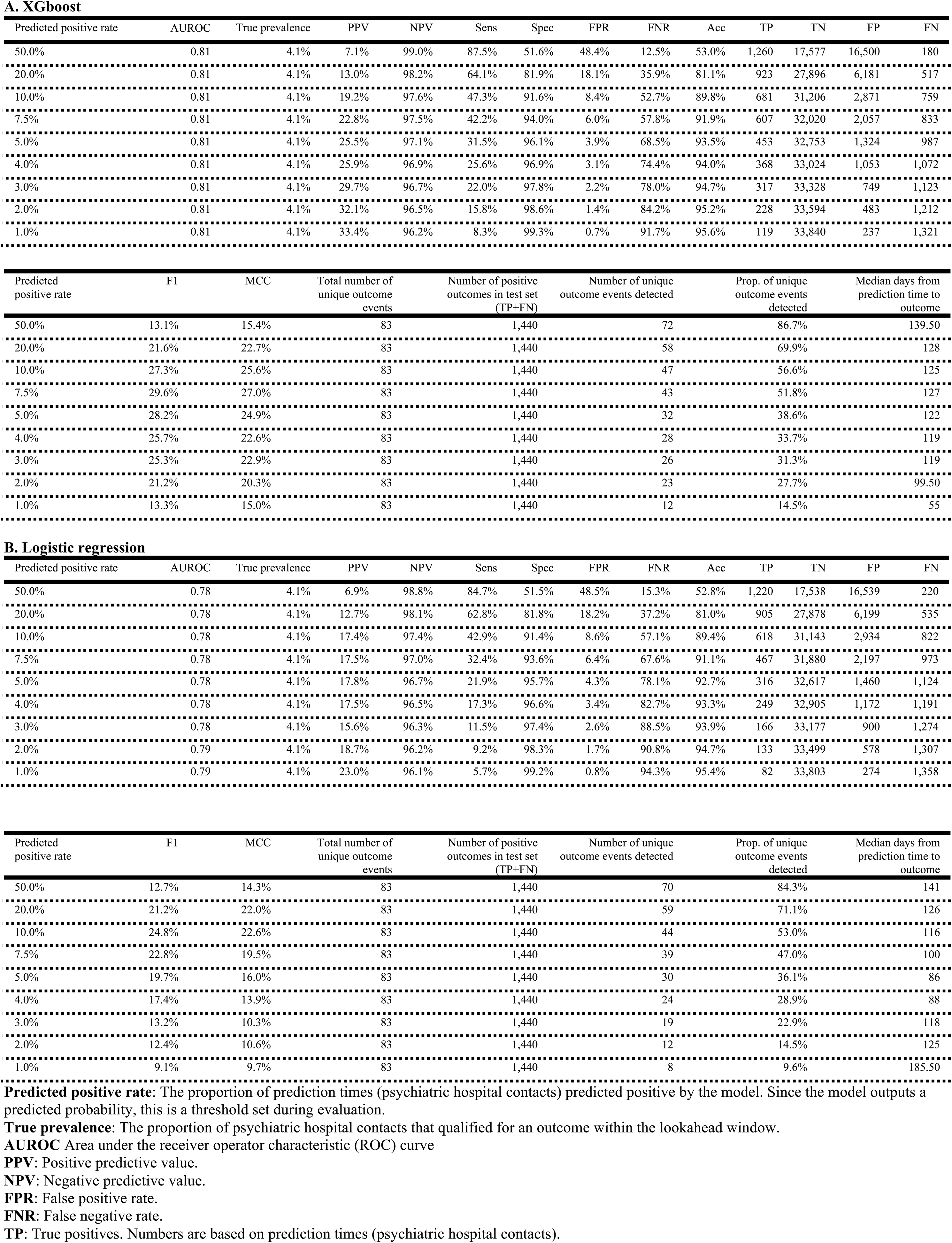
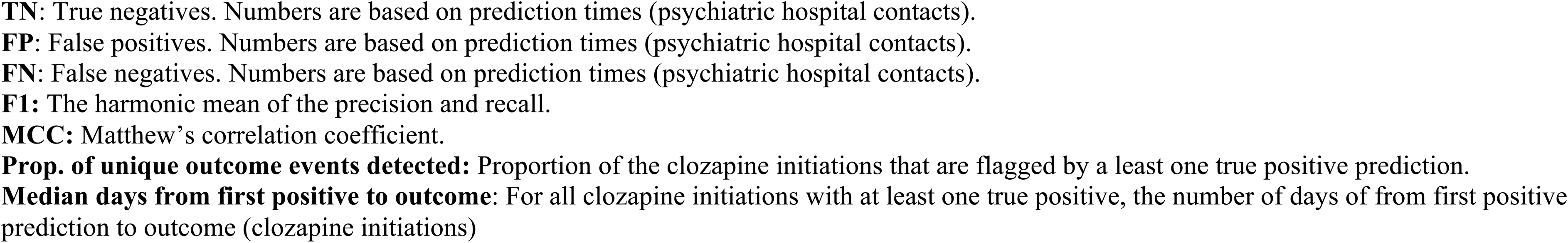
Performance metrics on test set for XGBoost and logistric regression model trained on full predictor set at varying positive rates.

Table 3 lists the performance metrics from the XGBoost (3A) and logistic regression (3B) model on the test set based on different predicted positive rates. At a PPR of 7.5%, the XGBoost model has a sensitivity of 42% and a positive predictive value of 23%. Thus, approximately two out of five of all true positive outcomes are correctly predicted, and for every five positive predictions, more than one prediction time is followed by a clozapine prescription within 365 days. At this PPR, 52% of the unique clozapine prescriptions are correctly predicted (true positive) at least once and the median time from first true positive to the clozapine prescription was 127 days. In comparison, at a PPR of 7.5%, the logistic regression model has a sensitivity of 32% and a positive predictive value of 18%. At this PPR, 47% of the unique clozapine prescriptions are correctly predicted (true positive) at least once and the median time from first true positive to the clozapine prescription was 100 days. Decision curve analysis of the models showed that both yield a universally greater net benefit than competing strategies in a sensible threshold probability range of 0.02 to 0.20. (see Supplementary Figure 8)

Supplementary Figure 4 and 5 shows the performance of the models across different patient characteristics, calendar time subgroups, and days to outcome. The models appear robust across all characteristics and the minor fluctuations, such as the variation in performance between age groups, can likely be attributed to similar minor differences in sample distributions. The sensitivity curves appear to remain stable as the time to outcome increases for both models. The calibration curves (see Supplementary Figures 6 and 7) indicate acceptable calibration for both models. Consistent with this, both models achieved similar Brier scores (XGboost: 0.0355 and logistic regression: 0.0379), modestly outperforming a prevalence-based null model (0.0393). The plots are cut off at predicted probabilities above 20% as there are too few patients with higher probabilities to make stable calibration estimates above this threshold.

Supplementary Table 6 and 7 lists the 30 predictors with the highest information gain (XGBoost) and standard coefficients (Logistic regression). In the interest of brevity, we focus on the best performing model (XGBoost – full predictor set). Here, 21 out of the 30 top predictors were text predictors. The TF-IDF predictors were based on the following terms from free text: “Tells”, “Suffering”, ”Hearing voices“, ”Expresses”, “VKO” (A Danish clinical acronym for objectively describing the patient as awake, clear and oriented), “Pain”, “Self-harm”, “Play”, “Voices”, “Deterioration”, “obs” (A Danish clinical term for a highlighting a clinical suspicion), “Headache”, “Family”, “Start”, “15 mg”, “Move”, “Increase”, “The voices”, “Normally”, “Depot” (likely referring to long acting injectable antipsychotics) and, “Neither”. The 11 remaining predictors were distributed on the following patient descriptors: Unique antipsychotics and antidepressants, benzodiazepines, anxiolytics, hospital contacts (both admissions and outpatient visits) to psychiatry, diagnosis of child and adolescent disorders (F9-subchapter of the ICD-10), Brøset Violence Checklist Score, and plasma risperidone.

### Post hoc analysis

After removing the text predictors “clozapine” and “leponex” from the full predictor set and retraining the models, XGBoost obtained an AUROC of 0.78 (5-fold interval: 0.76-0.80) and logistic regression an AUROC of 0.76 (5-fold interval:0.72-0.79) on the training data and similar results on the test set (XGBoost=0.79 and logistic regression=0.79).

## Discussion

This study developed and validated a dynamic prediction model for clozapine initiation using machine learning trained solely on EHR data from routine clinical practice. Specifically, predicting clozapine initiation at every outpatient visit to psychiatric services, the best model from the training phase achieved an AUROC 0.81 on the test set, flagging 42% of patients initiating clozapine initiation in the next year at least once with with a PPV of 23% at a positive predicted rate of 7.5. The model was well-calibrated and generally stable across demographics.

To our knowledge, this is the first dynamic prediction model for clozapine initiation trained on EHR data. This secures an alignment with the evolving nature of the morbidity of schizophrenia and schizoaffective disorder over the course of illness. Prior studies have employed a static approach, making predictions at index diagnosis or at the start of a treatment course.^4–6^ While this approach could provide clinicians with an early prediction of treatment resistance, it does not factor in clinical development over time, which is a substantial limitation. Compared to models developed using the static approach, our model demonstrated a higher predictive performance than comparable models, achieving an AUROC (or C-statistic for survival models) of 0.81 versus 0.69^4^, 0.70^5^ and 0.72^6^. These prior studies ^4–6^ were based on first episode psychosis (FEP) cohorts, whereas our model included patients irrespective of their stage of illness/time in treatment, including both incident cases and prevalent/chronic cases. This broader inclusion increases the generalisability of our model. Regarding the outcome definition, i.e., clozapine initiation, Osimo et al.^5^ used the same definition as in the present study. In contrast, Lee et al^6^ and Farooq et al^4^ defined treatment resistance through a lack of reduction in symptoms assessed on the Positive and Negative Syndrome Scale (PANSS30)^6^ or the Schedules for Clinical Assessment in Neuropsychiatry (SCAN)^4^ despite one period^6^ or two consecutive periods^4^ with antipsychotic drug treatment of adequate dose and duration. While these definitions of treatment resistance are, as opposed to the proxy used in the present study, arguably, more fine-grained and better aligned with the conventional definition of treatment resistance,^36^ they rely on symptom measures that are, unfortunately, not routinely implemented in clinical practice.^37^

To our knowledge, this is the first prediction model for treatment-resistant schizophrenia that applies natural language processing to generate predictors from clinical notes. Although the models that incorporated the full predictor set achieved the highest AUROC, those that relied only on TF--IDF predictors showed almost comparable performance. In the full predictor- model, the top features included TF--IDF predictors such as “hearing voices”, “suffering”, “selfharm-”, “depot” and “deterioration”, suggesting that this approach captures clinically significant constructs from the notes. However, other predictors such as “tells”, “play”, “start” and “move” lack sufficient context to be meaningfully interpreted. This illustrates a limitation of TF-IDF because it cannot represent- word context, a capability that more recent sentence transformer models provide. Conversely, sentence transformer models often sacrifice interpretability, while TF--IDF retains it. Notably in this regard, clinicians and legislators find explainability to be an important foundation for trust in tools that are considered for clinical implementation. ^38–40^

Regarding clinical utility, our model was relatively well calibrated and stable across demographic groups. It demonstrated a higher net benefit than both “treat all” and “treat none” strategies across all thresholds. Combined with its relatively high discriminative power, this suggests that the model may be suitable for implementation. If the prediction model were to be implemented as a clinical decision support- tool, a three-step approach for successful implementation has been proposed.^41^ First, the model runs silently on prospective EHR data without issuing alerts, allowing assessment of temporal stability, calibration, and early-stage error correction over a period matched to the incidence of clozapine initiation (typically 3–12 months). Second, if the silent phase demonstrates reliable performance, a targeted pilot is launched in a limited subset of outpatient clinics where intensive staff training, clear user guides, and close communication with local “champions” ensure proper adoption. Finally, the tool would be rolled out across psychiatric departments in the entire region with continuous monitoring of accuracy, drift, and subgroup bias.^41^

There are limitations to this study, which should be considered. First, clozapine prescription is a suboptimal proxy for treatment resistance because it captures only cases in which clozapine is actually prescribed. Clozapine treatment requires routine laboratory monitoring and high medication adherence, which some patients are unable to comply with. Consequently, the proxy misses cases of treatment-resistant schizophrenia. Second, the dataset included a relatively small number of outcomes and a relatively high number of predictors. This increases the risk of overfitting.^42^ However, several strategies were employed to reduce this. Specifically, structured predictors were selected based on the literature and clinical domain knowledge, and performance of the model was evaluated after 5-fold cross-validation. Additionally, the final evaluation was done on a hold-out test-set.^43^ Third, our model predicted the initiation of clozapine (intention-to-treat) and not the effect of clozapine treatment. Predicting who would benefit from clozapine treatment would be the ideal scenario. Studies show that 30-60% of those receiving clozapine respond.^44^ We do, however, not have the data on treatment effect as PANSS-30 or other symptom measures are not routinely collected. Fourth, the model was designed for the Central Denmark Region and if it were to be transferred to another region or country, re-tuning or re-training would be needed. Fifth, when using clinical notes as predictors, the resulting model might capture clinicians’ considerations regarding clozapine treatment prior to prescription. In such cases, the prediction model would merely confirm the clinician’s decision rather than adding new information. Because the 750 TF-IDF predictors included the terms “clozapine” and “leponex” (the Danish trade name for clozapine), we evaluated this issue by training and testing the models without these terms as predictors. This did, however, not reduce predictive ability substantially, indicating that prediction is not driven by mentions of the name of the drug in the clinical notes. Moreover, the primary models (including “clozapine” and “leponex” as TF-IDF terms) predicted clozapine initiation with a median lead time of approximately 127 days (see Table 3), suggesting that the models were not simply echoing clinicians’ notes just prior to the prescription.

In conclusion, we developed a dynamic prediction model solely based on EHR data from routine clinical practice, which predicts clozapine initiation with high discrimination. If implemented as a clinical decision support tool, this model may guide clinicians towards more timely clozapine initiation among patients with treatment-resistant schizophrenia.

## Data Availability

According to Danish law, the patient-level data for this study cannot be shared. The code for all analyses is available at: https://github.com/Aarhus-Psychiatry-Research/psycop-common/tree/main/psycop/projects/clozapine

## Acknowledgments

The authors are grateful to Bettina Nørremark for data management.

## Contributors

The contributors are reported according to CRediT. ^45^

Conceptualisation: EP, JGD, AAD, SDØ. Data curation and formal analysis: EP, JGD. Funding acquisition: SDØ. Investigation: Not relevant. Methodology: EP, JGD, AAD, SDØ. Project administration: EP, SDØ. Resources: EP, JGD, AAD, SDØ. Software: EP, JGD. Supervision: AAD, SDØ. Validation: EP, JGD. Visualization: EP, JGD. Writing – Original draft: EP. Writing – review & editing: EP, JGD, AAD, SDØ.

## Open Science

No pre-registration of the study was carried out. According to Danish law, the patient-level data for this study cannot be shared. The code for all analyses is available at: https://github.com/Aarhus-Psychiatry-Research/psycop-common/tree/main/psycop/projects/clozapine

