## Supplementary material for "Predicting clozapine initiation among patients with schizophrenia via machine learning trained on electronic health record data"

**TABLE OF CONTENTS**

**TABLES**

**Supplementary Table 1: Strategies for handling cases of doubt in manual text classification of clozapine initiation**

**Supplementary Table 2: Overview over patient descriptors and text embeddings/features used for predictor engineering**

**Supplementary Table 3: EHR clinical note types used for text features/embeddings**

**Supplementary Table 4: Hyperparameter tuning options during model training**

**Supplementary Table 5: Hyperparameters from the best XGboost and Logistic regression models during cross-validation**

**Supplementary Table 6: Information gain of top 30 model predictors from best XGboost model (full predictor set - 1 year lookahead))**

**Supplementary Table 7: Standardised coefficients of top 30 model predictors from best logistic regression model (full predictor set)**

**Supplementary Table 8: TRIPOD+AI checklist**

**FIGURES**

**Supplementary Figure 1: Cohort flowchart**

**Supplementary Figure 2: Histogram over timestamp differences between manual text validation and structured medicine data**

**Supplementary Figure 3: Histogram over outcome timestamps 2013-2024**

**Supplementary figure 4: Robustness of best XGboost model based on age, sex, month and days to outcome.**

**Supplementary figure 5: Robustness of best XGboost model based on age, sex, month and days to outcome.**

**Supplementary Figure 6: Plots of calibration and distribution of predicted probabilities for the XGboost model**

**Supplementary Figure 7: Plots of calibration and distribution of predicted probabilities for the logistic regression model**

**Supplementary Figure 8: Decision curve analysis plot for both XGBoost and Logistic regression model**

**Supplementary Table 1: Strategies for unsure cases in manual text validation**

|  | Description | Solution |
| --- | --- | --- |
| Historical prescription | Patients with documented prior clozapine treatment in the medical record, for example, “The patient was treated with clozapine between 2005 and 2010.” | Since the goal was to obtain data comparable to the prescription data, we did not exclude these as prevalent cases, as they would appear as incident cases in the period where prescription data were used to define clozapine initiation. |
| False incidents | Patients whose first documentation of clozapine treatment in the medical record occurred after January 1, 2014, but for whom it was evident that they were receiving continuous treatment. | These patients are considered prevalent users and were therefore excluded during cohort filtration. Data artifacts, such as incorrect timestamps in the notes, may cause patients to appear as prevalent users in the middle of the dataset. |
| Note clustering | Multiple notes share the same timestamp, suggesting they were administratively clustered, yet they describe clozapine treatment over time. For example, one note may state, “Rp Clozapine 25 mg,” while another, with the same timestamp, indicates, “Currently titrating clozapine, now at 150 mg.” | These cases were retained, and the first documented date was considered the incident date. |

**Supplementary Table 2: Overview of patient descriptors and text embeddings/features used for predictor engineering**

| **Group** | **Patient descriptor** | **Lookbehind (days)** | **Aggregation** | **N** |
| --- | --- | --- | --- | --- |
| **Static** | Sex (male/female)  Age (years) | - | - | 2 |
| **Hospital contacts** | All contacts  Psychiatric contacts  Somatic contacts | 30, 180, 365 | Count | 6 |
|  | Emergency contacts  Psychiatric contacts  Somatic contacts | 30, 180, 365 | Count | 6 |
|  | Admissions  Psychiatric admissions  Somatic admissions | 30, 180, 365 | Count, sum of hours | 12 |
| **Diagnoses (ICD-10)** | F0: Organic mental disorders  F1: Mental and behavioural disorders due to psychoactive substance use  F3: Mood/affective disorders  F30-F31: Manic and bipolar disorders  F4: Neurotic, stress-related, and somatoform disorders  F5: Behavioural syndromes associated with physiological disturbances and physical factors  F6: Disorders of adult personality and behaviour  F60.2-60.4 Cluster B personality (dissocial-, borderline- and histrionic personality disorder)  F7: Mental retardation  F8: Disorders of psychological development  F9: Behavioural and emotional disorders with onset usually occurring in childhood and adolescence or unspecified mental disorder^a^ | 30, 180, 365 | Boolean | 33 |
| **Medication** | Antipsychotics  1.generation  2. Generation  Olanzapine  Depot antipsychotics  Olanzapine  Aripiprazole  Risperidone  Paliperidone  Haloperidol  Perphenazine  Zuclopenthixol  Anxiolytics  Hypnotics  Sedatives  Antidepressants  Lithium  Alcohol dependence medications  Opioid dependence medications  Nervous system stimulants  Analgesics  Benzodiazepines related sleeping agents | 30, 180, 365 | Boolean | 60 |
|  | Regular prescriptions  Antipsychotics  Antidepressants  Anxiolytics  Analgesics | 30, 180, 365 | Unique count | 12 |
| **Coercive measures** | Involuntary admission and detention  Compulsory treatment  Involuntary medication  Electroconvulsive therapy  Involuntary treatment of somatic illness  Physical force  Manual restraint  Mechanical restraint  Mechanical restraint with straps  Chemical restraint | 30, 180, 365 | Mean**^b^** | 9 |
| **Psychometric rating scales** | Brøset Violence Checklist score  Suicide Risk Assessment | 30, 180, 365 | Mean, Latest | 12 |
| **Lab results** | Plasma-olanzapine  Plasma-aripiprazole  Plasma-risperidone  Plasma-paliperidone  Plasma-haloperidol  Plasma-paracetamol  Plasma-ethanol  Cancelled lab tests | 30, 180, 365 | Boolean | 24 |
| **Electroconvulsive treatment** | Electroconvulsive treatment | 30,180,365 | Boolean | 3 |
| **Text features/embeddings** | Term-frequency Inverse document frequency features | 180 | Notes within 180 days from a prediction time were concatenated to a single document and TF-IDF scores were calculated for the 750 uni- and bi-grams in the vocabulary. Se supplementary table 3 for selected notes) | 750 |

**^a^f99 is excluded ^b^Coercive measures is measured as duration.**

**Supplementary Table 3: EHR clinical note types used for text features/embeddings**

**EHR clinical note types**

Subjective mental state

Subjective physical state

Current objective mental state

Current objective physical state

Current social functioning

Semistructured diagnostic interview

Observation of patient

Reason for contact

Telephone consultation note

Appointments

Consultation with a Treatment Objective

Conclusion/final assessment

Plan

Order

Medicine

Psychiatric conference

Alcohol and substance use

**Supplementary Table 3: Hyperparameter tuning options during model training**

| **Preprocessing** |  |
| --- | --- |
| Imputation method for predictors with no data | Most frequent value, mean, median or no  imputation (only possible for XGBoost). |
| Scaling | Standard scaler or no scaling |
| **Model hyperparameters** |  |
| **XGBoost** |  |
| N estimators | [100; 1200] |
| Alpha | [10^-8^; 0.1] |
| Lambda | [10^-8^; 1.0] |
| Max depth | [3; 8] |
| Learning rate | [10^-8^; 1] |
| Gamma | [10^-8^; 10^-3^] |
| Grow policy | Either depthwise or lossguide |
| **Logistic regression** |  |
| Penalty solver | Elasticnet SAGA |
| C | [10^-8^; 10] |
| L1 ratio | [0; 1] |

**Supplementary Table 4: Hyperparameters from the best XGboost and logistic regression models during cross-validation**

|  | Model hyperarameters | | | | | | | | |
| --- | --- | --- | --- | --- | --- | --- | --- | --- | --- |
| **Model** | **Imputation** | **Scaling** | **N estimators** | **Alpha** | **Lambda** | **Max deptha** | **Learning rate** | **Gamma** | **Grow policy** |
| XGboost | Mean | Standard scaler | 293 | 6.75e^-05^ | 0.0163 | 4 | 0.033 | 1.55e^-06^ | depthwise |
|  |  |  | **tol** | **C** | **Intercept scaling** | **Random state** | **Penalty solver** | **Max iter** | **L1-ratio** |
| Logistic regression | Median | Standard scaler | 0.0001 | 0.00378 | Fit intercept | 41 | Elastic net saga | 134 | 0.56 |

**Supplementary Table 5: Information gain of top 30 model predictors from best XGboost model (full predictor set – 1 year lookahead)**

| **LOOKBEHIND WINDOW** | AGGREGATION FUNCTION | PREDICTOR^1^ | INFORMATION GAIN |
| --- | --- | --- | --- |
| **365** | Unique count | Antipsychotics | 0.007 |
| **180** | Bool | Benzodiazepines | 0.006 |
| **30** | Bool | Anxiolytics | 0.006 |
| **180** | TF-IDF | “Tells” | 0.005 |
| **180** | TF-IDF | ”Suffering” | 0.005 |
| **180** | TF-IDF | “Hearing voices” | 0.005 |
| **180** | TF-IDF | ”Expresses” | 0.005 |
| **365** | Unique Count | Antidepressants | 0.005 |
| **180** | TF-IDF | ”VKO”^2^ | 0.005 |
| **180** | TF-IDF | ”Pain” | 0.004 |
| **180** | TF-IDF | ”Self-harm” | 0.004 |
| **185** | TF-IDF | ”Play” | 0.004 |
| **30** | Count | Physical visits to psychiatry | 0.004 |
| **180** | TF-IDF | “Voices” | 0.004 |
| **180** | Bool | Diagnosis of child and adolescent disorder^3^ | 0.004 |
| **180** | TF-IDF | ”Deterioration” | 0.004 |
| **180** | TF-IDF | ”obs”^4^ | 0.004 |
| **180** | TF-IDF | ”Headache” | 0.004 |
| **180** | TF-IDF | ”Family” | 0.003 |
| **180** | TF-IDF | ”Start” | 0.003 |
| **180** | TF-IDF | ”15 mg” | 0.003 |
| **180** | TF-IDF | ”Move” | 0.003 |
| **180** | Unique count | Antipsychotics | 0.003 |
| **180** | TF-IDF | ”Increase” | 0.003 |
| **180** | TF-IDF | ”The voices” | 0.003 |
| **30** | Mean | Brøset Violence Checklist Score | 0.003 |
| **365** | Bool | Plasma risperidone | 0.003 |
| **180** | TF-IDF | “Normally” | 0.003 |
| **180** | TF-IDF | ”Depot” | 0.003 |
| **180** | TF-IDF | ”Neither” | 0.003 |

^1^TF-IDF predictor labels are translated from Danish. ^2^ “VKO” is a Danish clinical acronym for objectively describing the patient as awake, clear and oriented, ^3^ F9-subchapter of the ICD-10. F99 was not included in this group. ^4^ “obs” = A Danish clinical term for a highlighting a clinical suspicion.

**Supplementary Table 6: Standardised coefficients of top 30 model predictors from best logistic regression model (full predictor set - 1 year lookahead)**

| **LOOKBEHIND WINDOW** | AGGREGATION FUNCTION | PREDICTOR^1^ | STANDARDISED COEFFICIENTS |
| --- | --- | --- | --- |
| **Top positive coefficients** | | | |
| **365** | Unique count | Antipsychotics | 0.33 |
| **180** | TF-IDF | ”Clozapine” | 0.17 |
| **180** | TF-IDF | ”The voices” | 0.16 |
| **180** | TF-IDF | ”Distressed” | 0.09 |
| **180** | Unique count | Antipsychotics | 0.13 |
| **180** | TF-IDF | ”Olanzapine” | 0.11 |
| **180** | TF-IDF | ”Effect” | 0.11 |
| **180** | TF-IDF | ”Sat” | 0.11 |
| **180** | TF-IDF | ”Week” | 0.11 |
| **365** | Unique count | Antidepressants | 0.11 |
| **365** | Boolean | Plasma-paliperidone | 0.11 |
| **365** | Boolean | Long-acting paliperidone | 0.10 |
| **180** | Count | Admission to a psychiatric hospital | 0.10 |
| **180** | TF-IDF | ”Leponex” | 0.10 |
| **180** | TF-IDF | ”Sentence”^2^ | 0.10 |
| **Top Negative Coefficients** | | | |
| **365** | Count | Emergency contacts to psychiatry | -0.26 |
|  | Static | Age | -0.15 |
| **180** | TF-IDF | ”Manage” | -0.14 |
| **180** | TF-IDF | ”Mood” | -0.08 |
| **180** | TF-IDF | ”Help” | -0.13 |
| **180** | TF-IDF | ”Contacts” | -0.11 |
| **180** | TF-IDF | ”Meets” | -0.11 |
| **365** | Latest | Suicide risk assessment | -0.11 |
| **180** | TF-IDF | ”Responds” | -0.11 |
| **180** | TF-IDF | ”Emotional” | -0.10 |
| **180** | Count | Emergency contacts due to a physical disease | -0.09 |
| **180** | TF-IDF | ”14” | -0.09 |
| **180** | TF-IDF | ”City” | -0.08 |
| **180** | TF-IDF | ”The admission” | -0.08 |
| **180** | TF-IDF | ”Municipality” | -0.08 |

^1^TF-IDF predictor labels are translated from Danish. ^2^ Refers to sentence to psychiatric treatment

**Supplementary Table 8: TRIPOD+AI checklist**

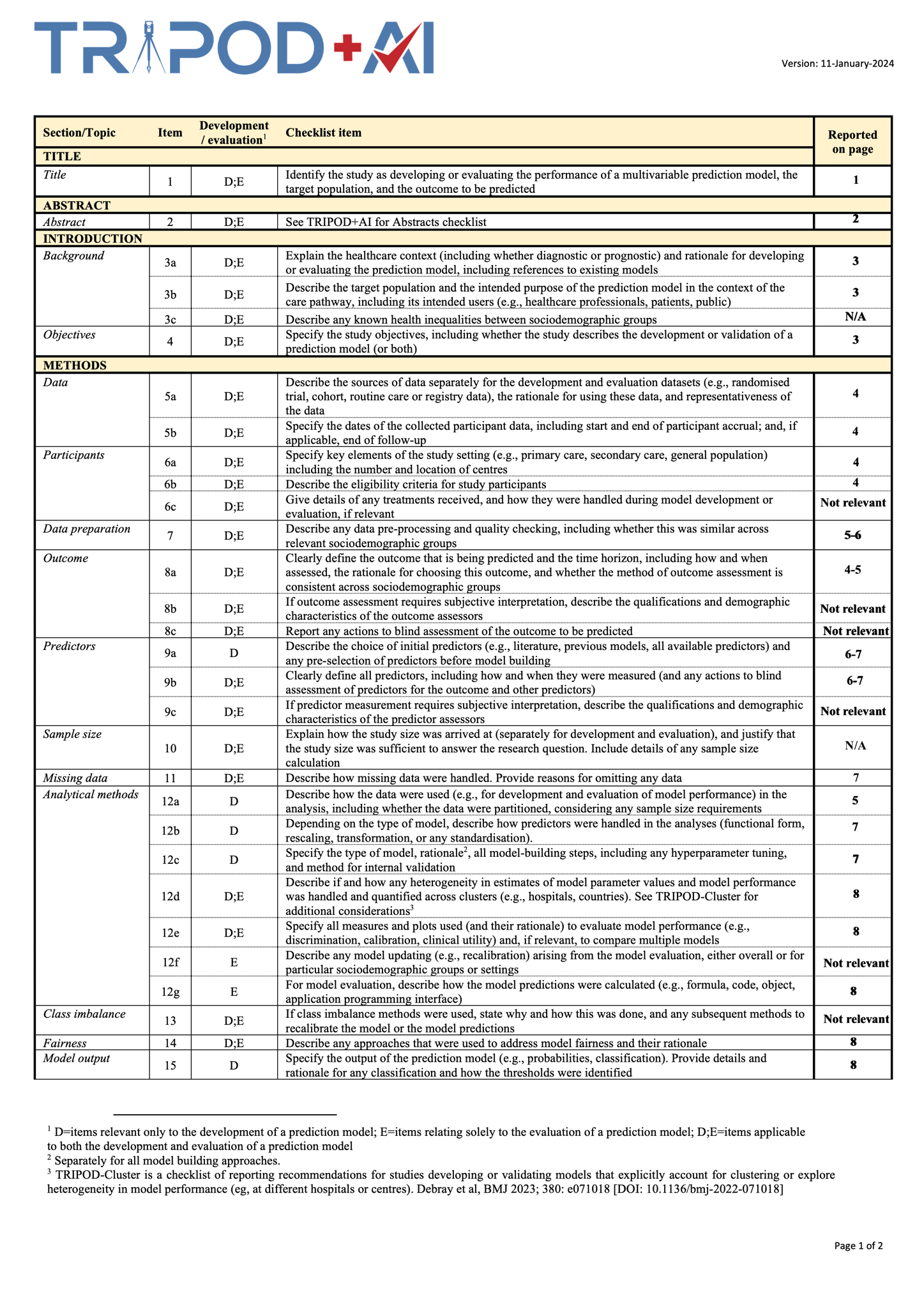

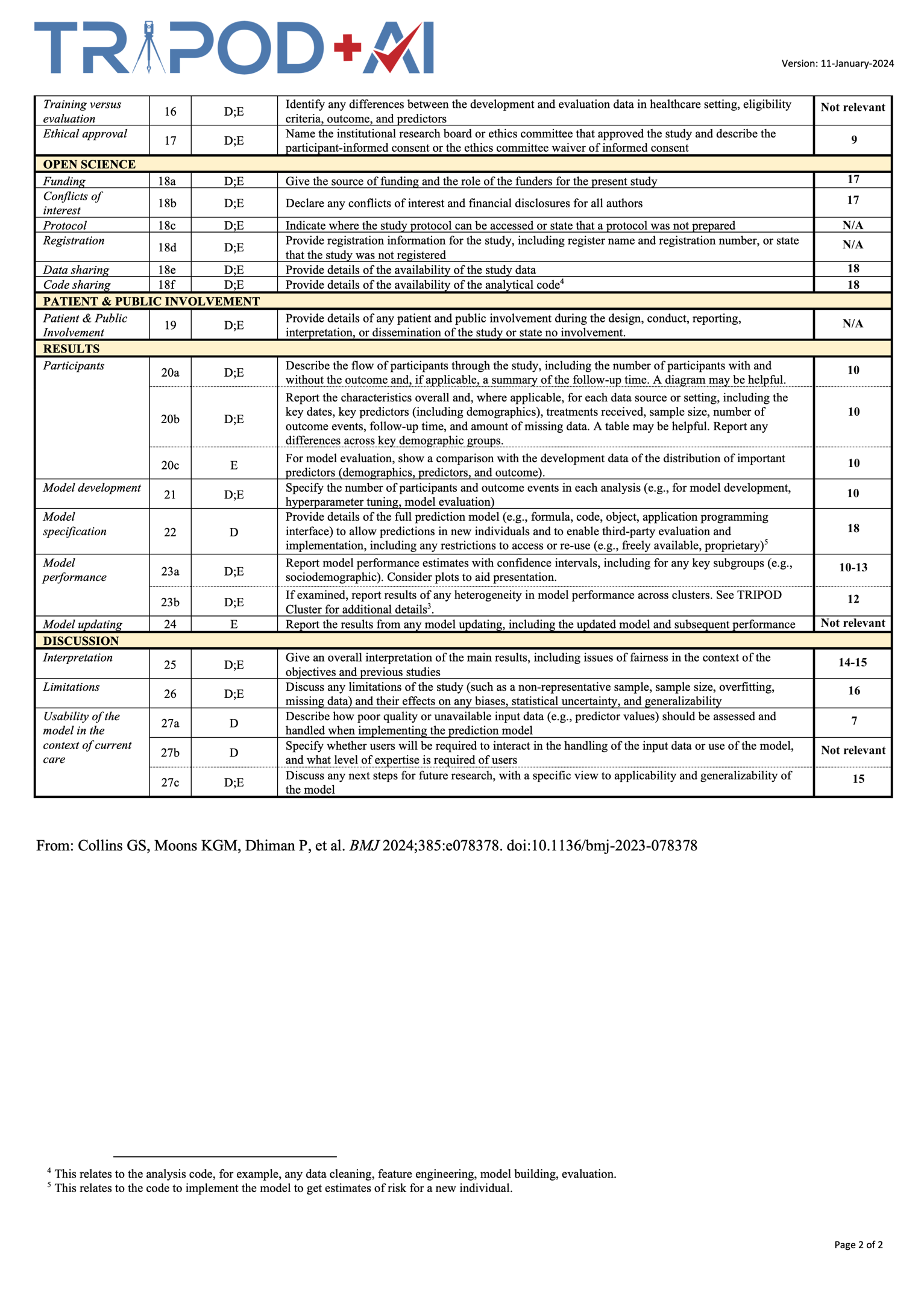

**FIGURES**

**Supplementary Figure 1: Cohort flowchart**
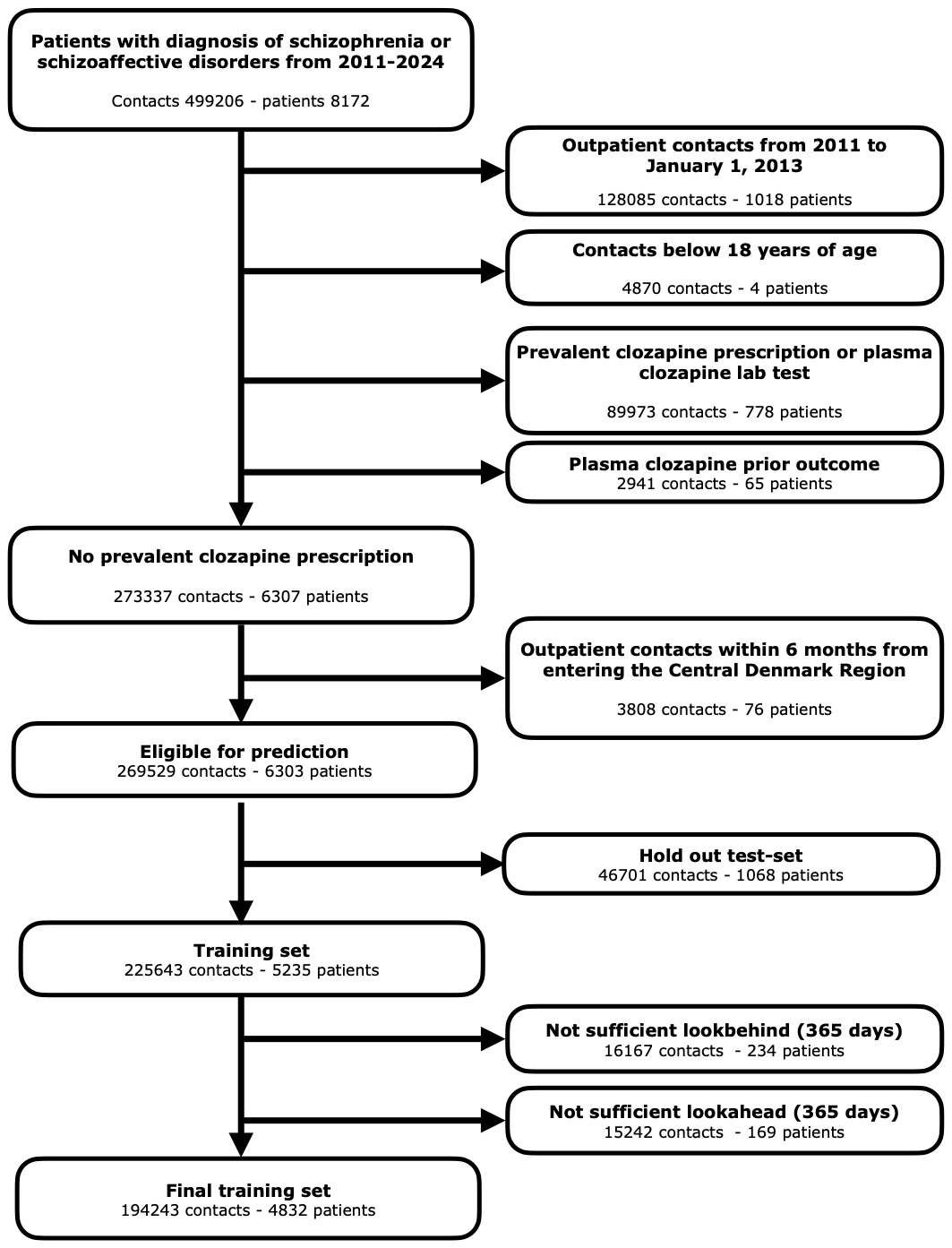

**Supplementary Figure 2: Histogram over timestamp differences between manual text validation and structured medicine data**

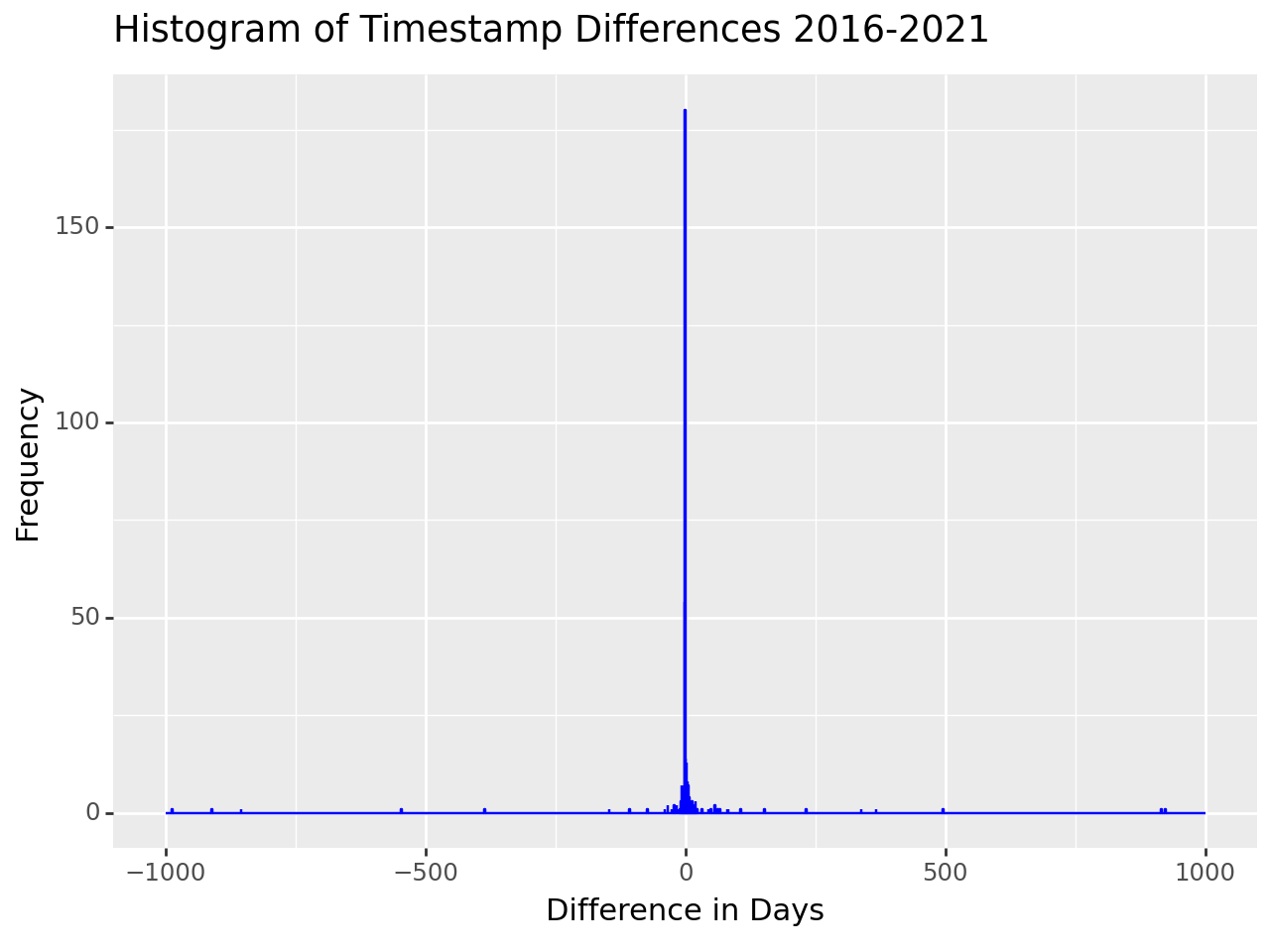

**Supplementary Figure 3: Histogram over outcome timestamps 2013-2024**

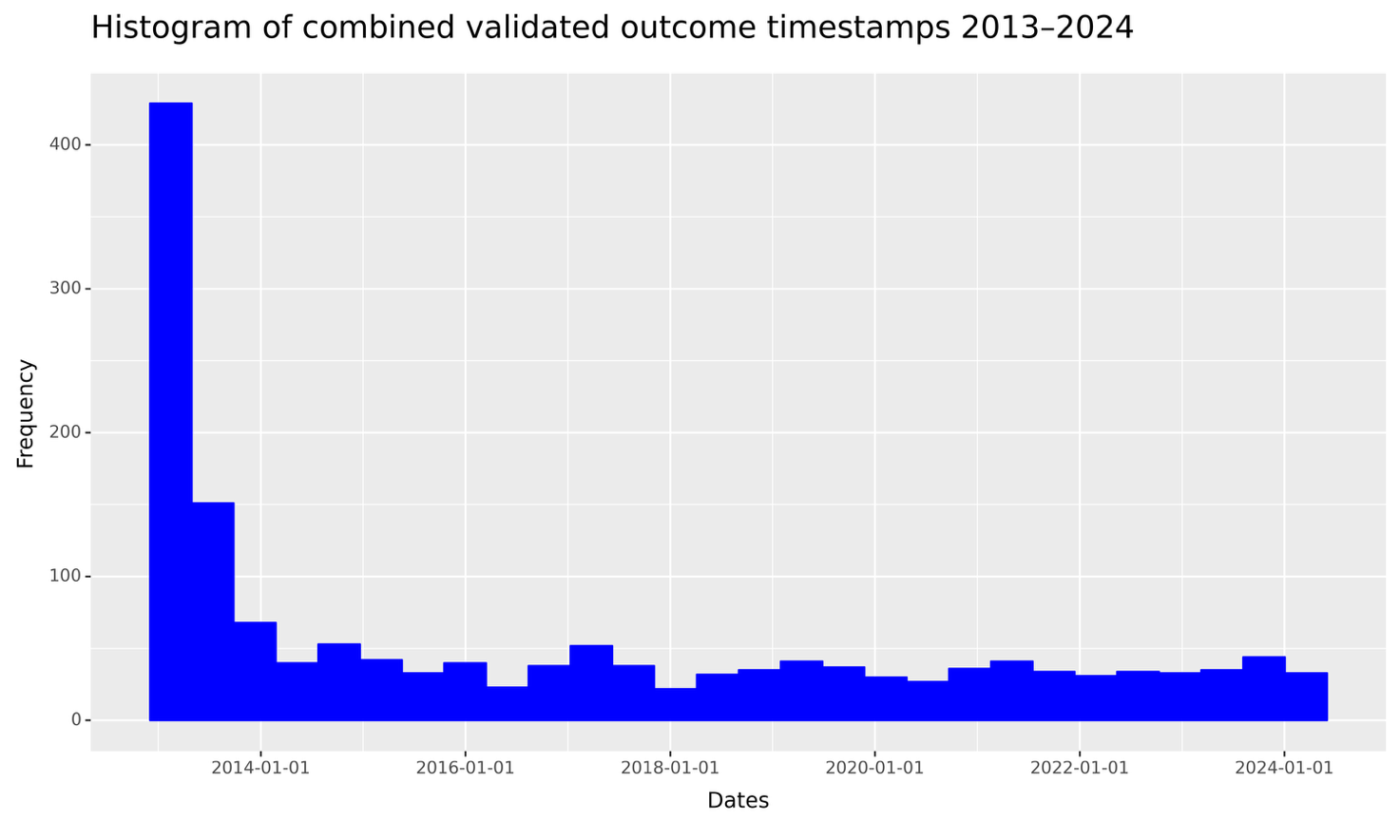

**Supplementary figure 4: Robustness of best XGboost model based on age, sex, month and days to outcome.**

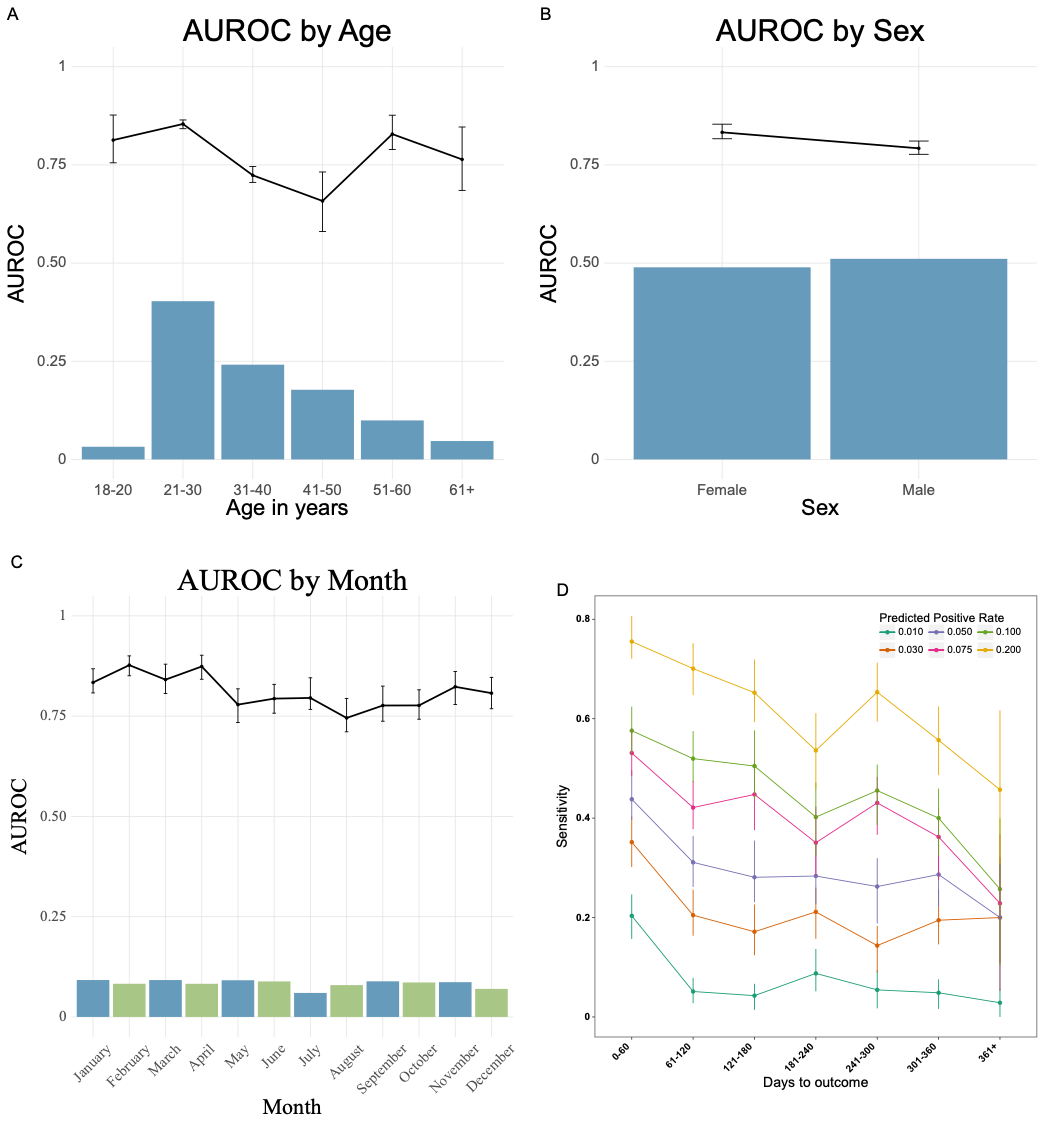

**Supplementary figure 5: Robustness of best logistic regression model based on age, sex, month and days to outcome.**

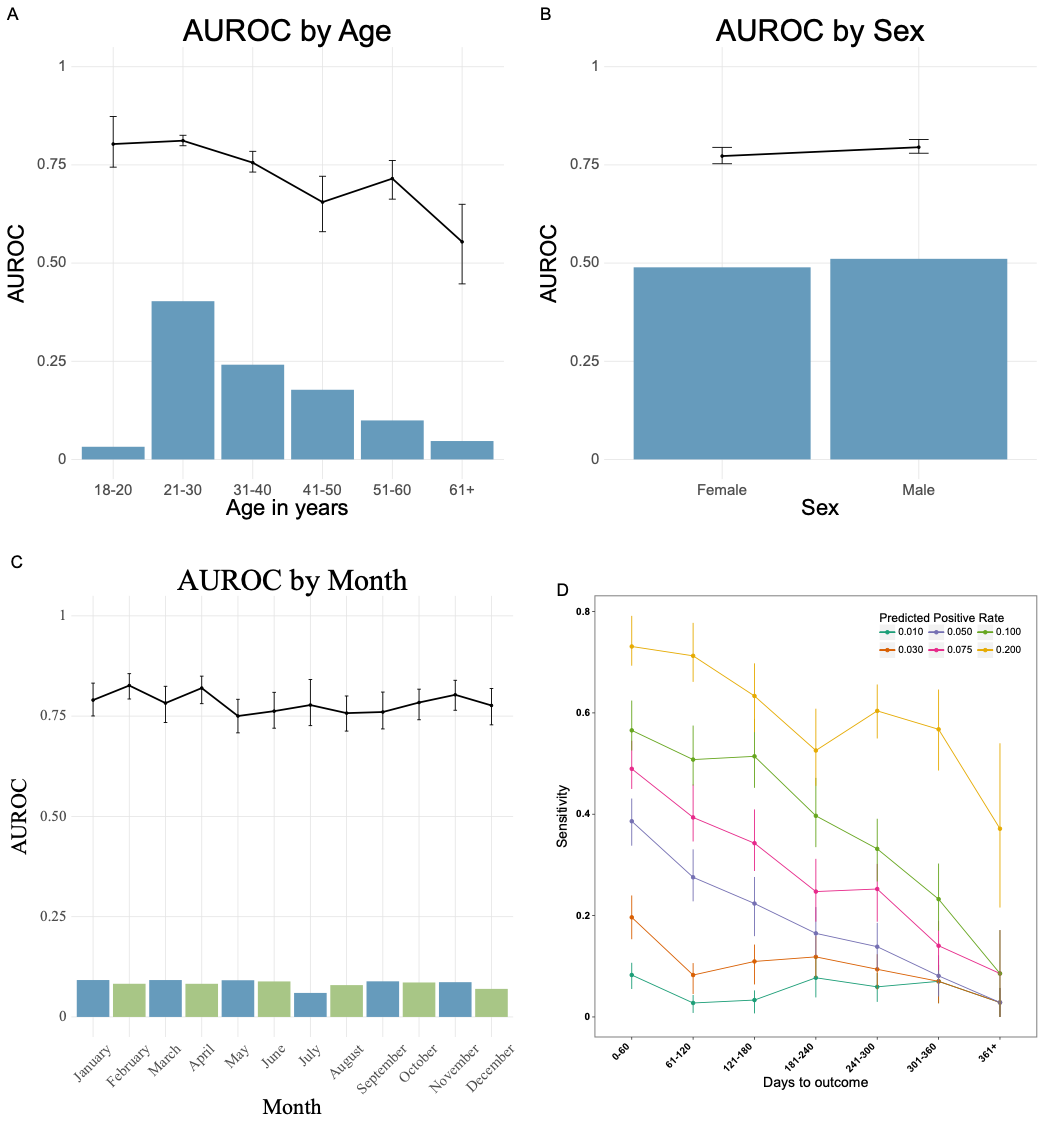

**Supplementary Figure 6: Plots of calibration and distribution of predicted probabilities for the XGboost model**

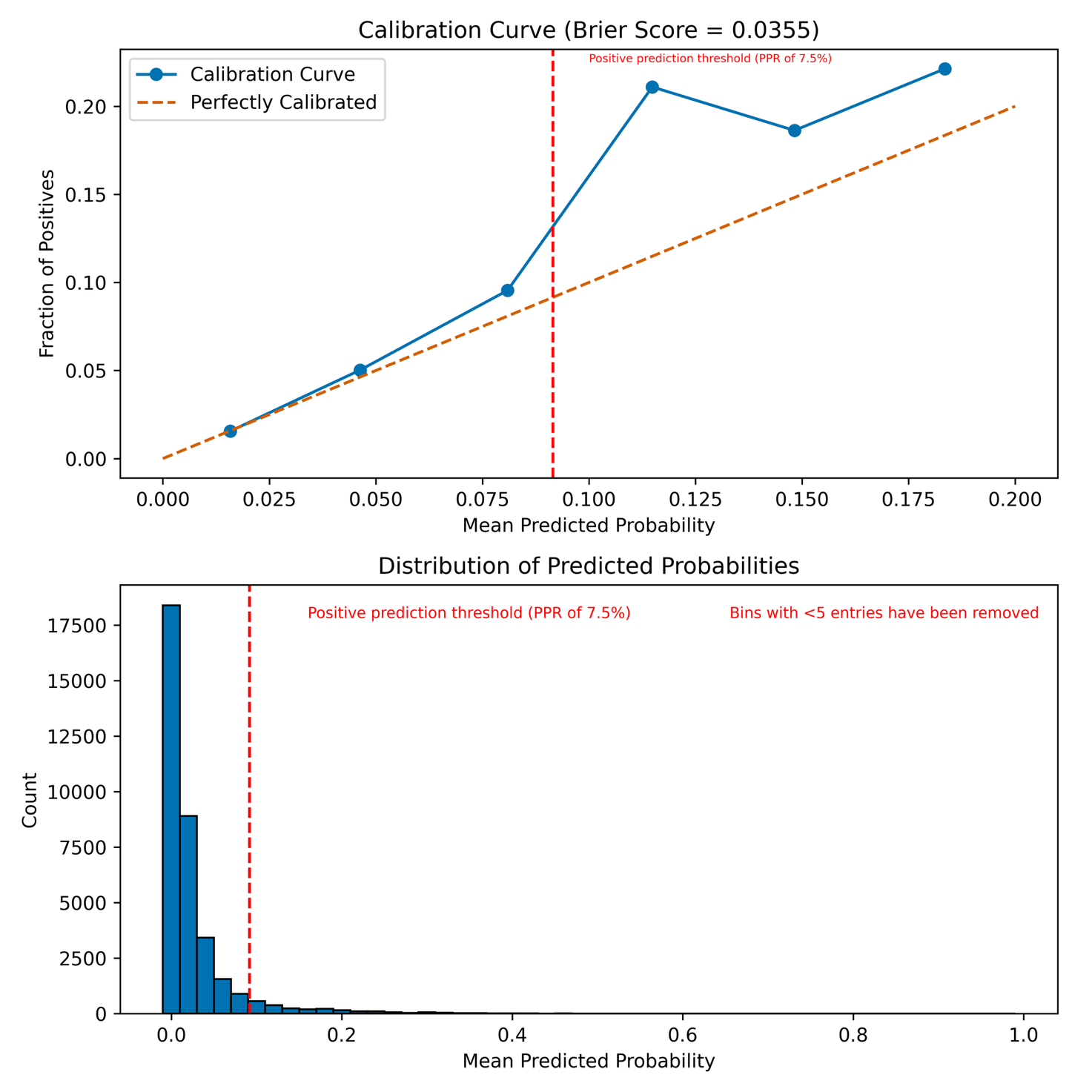

**Supplementary Figure 7: Plots of calibration and distribution of predicted probalities for the logistic regression model**

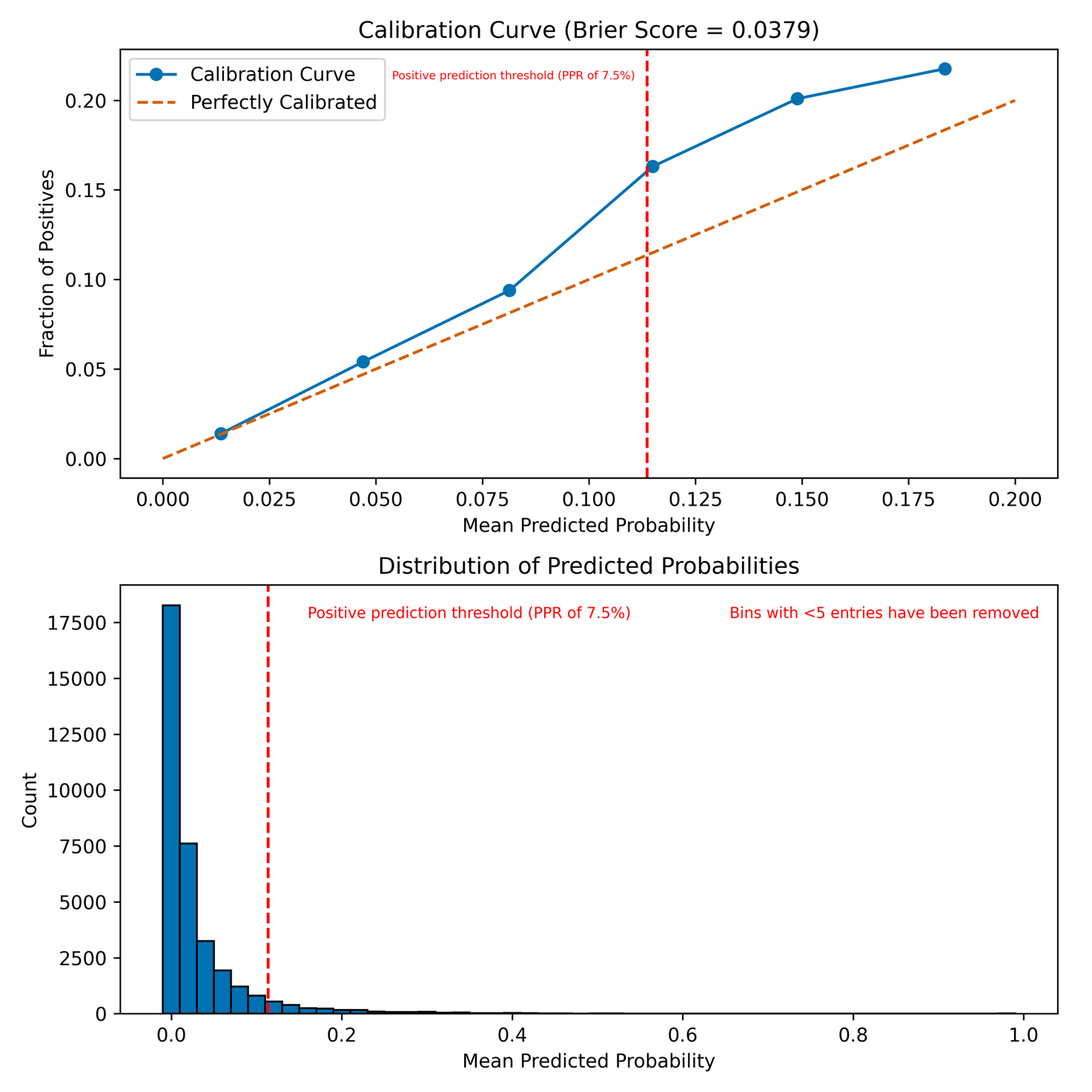

**Supplementary Figure 8: Decision curve analysis plot for both XGBoost and logistic regression model**

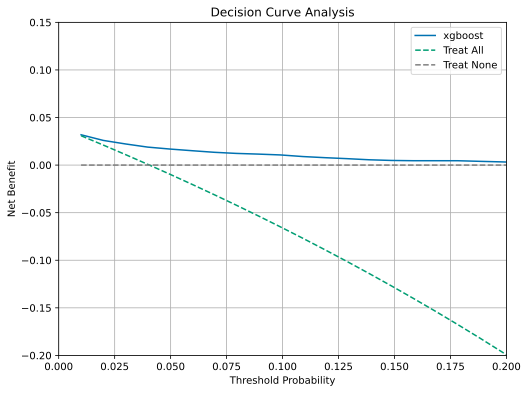

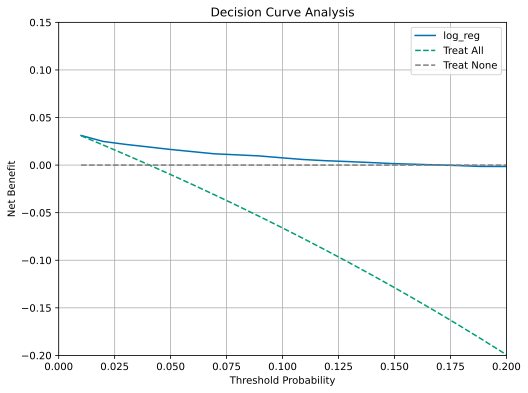
